# DALSO: domain ALS ontology

**DOI:** 10.1101/2024.06.01.24308128

**Authors:** Teresa Podsiadły-Marczykowska, Peter Andersen, Marta Gromicho, Julian Grosskreutz, Magdalena Kuźma-Kozakiewicz, Susanne Petri, Katarzyna Szacka, Hilmi Uysal, Mamede de Carvalho, Maria Piotrkiewicz

## Abstract

Amyotrophic lateral sclerosis (ALS) is an incurable, rapidly progressive neurodegenerative disease. During the course of ALS, virtually all skeletal muscles are gradually affected, including the respiratory muscles, and the disease is usually fatal within 2–5 years of symptom onset. Unequivocal and conclusive tests for ALS do not exist, its disease etiology is still unknown, and therapeutic options are limited. This paper presents the ALS domain ontology (DALSO), model containing formalized, semantic descriptions of a wide range of modeled disease related notions such as patient demographics, clinical findings and history, disease clinical features and diagnostic classifications, risk factors, genetics and pathophysiological mechanisms of motor neuron degeneration. The DALSO’s aim and information scope, design assumption, structure and implementation details are also described. DALSO covers the broad range of significant biomedical concepts ranging from clinical to molecular feature of the modeled disease, it represents a comprehensive, structured knowledge source for the ALS disease domain. To the best of authors’ knowledge, the DALSO is the first attempt to develop a formal, computational model representing knowledge of this fatal motor neuron disease. It provides the means for integrating and annotating clinical and research data, not only at the generic domain knowledge level, but also at the level of individual patient case studies. The DALSO is expressed in OWL2 language, contains 910 classes, is consistent and free of logical errors. Its syntactic correctness was validated by the Fact++ reasoner.

## 1. Introduction

### 1.1 ALS disease features

The etiology of amyotrophic lateral sclerosis (ALS) is still unknown, although this disease was first reported in 1869 by the French neurologist Jean-Martin Charcot (Charcot and Joffroy 1869). ALS is a fatal, rapidly progressive neurodegenerative disorder characterized by degeneration of both the upper motor neurons (MNs) of the motor cortex and lower MNs of the brainstem and spinal cord. The initial symptoms consist of progressive muscle weakness, wasting and spasticity. During the course of ALS, all voluntary muscles are eventually affected, including the respiratory muscles, and the disease is usually fatal within 2–5 years of symptom onset. There is a relatively small subgroup of patients that demonstrate slower disease progression, surviving even beyond 10 years (Witzel et al. 2022). Recently, the possibility was suggested that disease course becomes slower over time (Czaplinski et al. 2006). Respiratory insufficiency always occurs in patients with ALS, generally a the end stage, and is a major cause of mortality.

The mean age of onset is 60 years, but symptoms can start at any adult age. The male to female ratio is approximately 1.4:1, with hormones possibly playing a protective role in premenopausal women (Manjaly et al. 2010). The incidence of ALS is 2–4/100,000 per year, with heterogeneous distribution worldwide (Marin et al. 2017), and a prevalence of about 2– 5/100,000 (Chiò et al., 2013), possibly increasing over the years, which can partly be explained by lengthening lifespan (Korner et al. 2013). The life-time risk is in the order of 1/400 for males and 1/550 for females (Ryan et al. 2019).

Approximately 5% of ALS patients have a first-degree relative with ALS. When more distant relatives are taken into account, about 10% of patients are found to have a familial case of ALS (fALS), and the remaining 90% of ALS cases are apparently sporadic (sALS) (Mejzini et al. 2019). It has been shown that genetic factors are involved in the pathogenesis of around 70% of fALS cases. Moreover, mutations in the same genes were found in evidently sporadic cases, although at lower frequencies, which resulted in oligogenic hypothesis suggesting that sALS cases also may be caused by yet undiscovered genetic mutations (Kim et al. 2020; van Blitterswijk et al. 2012; Alsultan et al. 2016). This hypothesis was tested in recently published study aimed towards characterization of genetic variability in genes related to sALS (Van Daele et al. 2023). The authors studied genetic variation in 90 ALS-associated genes in a large cohort of 6529 sALS patients. The study confirmed genetic involvement only in 28.17% of patients, but concluded that further research to test oligogenic hypothesis is needed.

Therapeutic options for ALS are limited and no preventive treatment exists. The most important part of current patient care consists of nutritional and respiratory management and alleviation of symptoms. The only drugs currently approved for ALS treatment are riluzole and edaravone, both providing relatively modest effects. Moreover, positive effects of edaravone are limited to a specific subpopulation of patients.

Nevertheless, the search for effective ALS therapies continues around the world. First of all, we should mention the Institute for the Development of ALS Therapy, whose research is financed by the ALS community. Several other clinical trials of new treatments are also ongoing (e.g. (Miller et al. 2022; Wiesenfarth et al. 2024), see also https://www.eventalways.com/2nd-als-drug-development-summit and https://www.biopharmadive.com/news/als-drug-research-progress-genetic-clinical-trials/611274/ Hopefully these activities will soon result in the emergence of effective therapies for ALS, and also bring us closer to understanding the etiology of this disease.

A fact that can aggravate prognosis of ALS is diagnostic delay (Campos et al. 2021, 2023) averaging 5–27 months, depending on the disease onset region (Sennfält et al. 2023), which urge to be changed (Andersen et al. 2012; Segura et al. 2023). This can be primarily attributed to the clinical heterogeneity of ALS, recognized as a consistent disease feature (e.g. Goutman et al. 2022) due to the existence of many conditions mimicking ALS (Traynor et al. 2000) and to the very commonly subtle symptoms of ALS onset.

An unequivocal and conclusive test for ALS does not exist (Turner and Swash 2015), despite many years of research and undeniable progress in understanding the disease mechanisms. Currently, the ALS diagnostic process is primarily based on clinical assessment by an experienced neurologist, relying heavily on individual knowledge and experience, although diagnostic support through electrophysiological investigations, electromyography (Fukushima et al. 2022; Goutman et al. 2022) and neuroimaging (Cengiz and Kuruoğlu 2020; Verstraete and Foerster 2015) is also recommended.

Despite the huge body of research into genetics, epidemiology, diet, lifestyle, and the many mechanisms of cellular pathogenesis observable in ALS, a single unifying theory for the disease etiology is still lacking.

### 1.2 Motivation for the ALS domain ontology construction

Domain ontological models have two interrelated aspects. The first is a specialized domain terminology, the second represents semantics inherent to particular terms. Both model aspects are crucial to the realization of domain model applications such as unequivocal data capture, database schema construction, data integration (Calvanese et al. 2017b; Zhang et al. 2017) or, relevant to the genetic data analysis, extensions of enrichment analysis (Shah et al. 2012). Other important features of the ontological domain model are explicit taxonomy and modular structure, semantic explicitness, explicit linkage of concepts and relations, and the final expression of concept definitions using machine readable formal language. These features indicate that an ontological model allowing for the semantic encoding of domain terminology is the method of choice for the integration and annotation of heterogeneous data sources and it can also be helpful in the formulation of unambiguous diagnostic criteria (Abdollahi et al. 2021; Steiner et al. 2017).

These features of ontological models are reasons for which they have been proven as practical solutions in biomedical areas such as: (i) knowledge formalization and sharing (Sánchez and Moreno 2008; Staab and Studer 2010), (ii) logical, unequivocal concept definition in biomedicine (Bug et al. 2008; Silachan and Tantatsanawong 2011; Tao et al. 2013) (iii) integration, comparison and reuse of data and information (Ruttenberg et al. 2009; Ivanović and Budimac 2014; Nielsen 2014), and eventually (iv) new methods for genetic data analyses (Shah et al. 2012; Bean et al. 2020; Segura et al. 2023).

Besides the lack of a definitive test for diagnosing ALS, sound clinical assessment of ALS patients is additionally complicated by the inherent deficiency of the precision of terms describing the symptoms, diagnostics and phenotypic criteria. (Ruttenberg et al. 2009) state that there is no clear distinction between diagnostic and phenotypic ALS classifications because the vocabulary of terms used for clinical patient features, descriptions and classification of ALS phenotypes has often been inconsistent. The first three of the above-mentioned features of ontological models indicate that they have the potential to provide a solution to ALS diagnostic problems.

The genetic causes of ALS remain unclear today and a better understanding of patients’ genetic data could result in deeper insights into the mechanisms of the disease. This issue can also be addressed using ontological models.

Gene ontology (GO) (Tao et al. 2013), a widely accepted reference ontology in biomedicine, is today a valuable tool for describing gene functions, although it has known limitations, such as the lack of disease specific context. Therefore, GO cannot be used to perform specific disease-related enrichment analysis considering e.g. the problem of disease subtypes, syndromes or phenotypes, under-or over-represented in the given gene set in question (Osborne et al. 2009).

There are reference ontologies containing human disease-related concepts/terminology, but these models are characterized by high-level, broad coverage knowledge, which limits their use for specific disease problems. Shah et al. (2012) proposed the novel method of enrichment analyses consisting of referencing specific disease domain ontologies with GO.

In addition, further research on the cell-specific biochemistry and physiology of MNs and the cellular pathways disturbed during MN degeneration are likely to lead to the development of more effective neuroprotective treatments for patients.

In order to find possible created and published ALS domain ontology or ontologies related to ALS, we have searched internet resources, including PubMed, ScienceDirect, IEEE Xplore, Google Scholar, Research Gate, and Springer Link, along with the special purpose semantic search engine Watson and classical key word search using Google. Moreover, we searched also repositories such as the NCBO BioPortal (http://bioportal.bioontology.org/), currently the most comprehensive biomedical ontology repository, Ontology Lookup Service from the European Bioinformatics Institute (http://www.ebi.ac.uk/ontologylookup) and the Open Biomedical Ontology (OBO) Foundry (http://www.obofoundry.org/). The searches returned only one ALS-related ontology (OntoPaRON), which was used to describe the care pathway for patients from the Ile-de-France ALS network (Cardoso et al. 2021). OntoPaRON concentrates on the notions relating to area of medical care and social help for ALS patients, but the representation of medical concepts is poor and confined to basic anatomical concepts.

Therefore we concluded that to the best of our knowledge, the ontology of the ALS domain presented in this article is the first attempt to develop a semantic, computational model of ALS knowledge.

Notably, the search found two domain ontologies related to other neurodegenerative diseases, namely Parkinson’s (Younesi et al. 2015) and Alzheimer’s (Malhotra et al. 2014). In this way, the new ALS model will complement the spectrum of existing models related to neurodegenerative pathology.

The domain ontology of ALS disease should enable the integration of clinical and genetic data, as well as extended analysis of these data, which could ultimately translate into solving many problems both in ALS research and in clinical practice.

The paper is organized as follows: second section presents ontology construction methodology as well as knowledge acquisition and reuse problem; third section provides general model description, giving also in subsections the details of the ontology modules; section four concludes the paper.

## 2. DALSO construction – methodology, knowledge acquisition and reuse

DALSO development is closely related to the OnWebDUALS research project.

### 2.1. Ontology-based Web Database for Understanding ALS (OnWebDUALS)

The OnWebDUALS project was carried out under the EU Joint Programme – Neurodegenerative Disease Research (JPND) in 2015-2018. The project consortium consisted of 6 neurology departments from 5 countries, genetic laboratory from Sweden, and one research team from Nalecz Institute of Biocybernetics and Biomedical Engineering, Warsaw.

The main objective of this project was to investigate the interplay between the characteristics of ALS patients, their genetic profiles, and environmental influence. In addition, other aspects were investigated, such as the pattern of disease spreading from the first affected region to other regions of the body, family history, related diseases, injuries and surgeries, exercise, and diagnostic pathway in different countries.

The central to this project was to build an ALS domain ontology comprehensively representing a body of related medical knowledge and serving as the formal basis for building a standardized E-health record for anonymous ALS was patients, implemented in the European ALS Web-database.

ALS patients were interviewed during clinical practice using a standard questionnaire developed by the OnWebDUALS consortium at five European University Hospital centers: Akdeniz University, Antalya, Turkey; Hannover University Medical School, Germany; Jena University Hospital, Germany; Faculty of Medicine, University Hospital of Lisbon, Portugal; Medical University of Warsaw, Poland. Clinical evaluation at each center was performed by a designated senior neurologist with extensive clinical experience in the treatment of ALS. Ethical consent to implement the project was granted by the relevant Clinical Ethics Committees of the participating centers, and each patient and control subject gave written informed consent to the collection and analysis of his/hers medical data.

The recruitment process permitted to interview a total of 1376 patients from the following centers: University Hospital Jena: 148 patients; Hannover Medical School: 236 patients; Medical University of Warsaw: 286 patients; Faculty of Medicine, University of Lisbon: 471 patients; and Akdeniz University Faculty of Medicine, Antalya: 235 patients. For comparisons, about 1000 controls from the above referred centers were included.

Based on this large dataset, several papers were published so far, describing the influence of environment and lifestyle factors on ALS onset and progression (Kuraszkiewicz et al. 2018; Korner et al. 2019), risk factors for polyneuropathy in ALS (de Carvalho et al. 2020), the patterns of disease presentation (Prell et al. 2020; Gromicho et al. 2020; Gromicho et al. 2021; Matos et al. 2020), diagnostic pathway of ALS patients (Campos et al. 2021, 2023), the rate of disease progression (Prell, 2020; Barc et al. 2020), the possible biomarkers (Hertel et al. 2022), the impact of comorbidities (Diekmann et al. 2020; Pereira et al. 2021), the predictive impact of C9Orf72 mutation in respiratory decline (Miltenberger-Miltenyi et al. 2019), family history of neurodegenerative diseases (Campos et al., 2020), influence of ALS on fertility (Uysal et al. 2021), association of the contact sports with ALS (Henriques et al. 2023), as well as potential predictive measures (Kuraszkiewicz et al. 2020).

### 2.2 Applied methodology, DALSO aim and range

METHONTOLOGY (Fernández-López et al. 1997) is a method of ontology building that is grounded in software engineering methodologies. It is well suited for building ontologies from scratch, but also recommends and supports ontological knowledge reuse.

The main, general aim of DALSO is to formalize medical terminology in the domain of ALS disease, embedding it in the broad context of knowledge involved in ALS diagnosis, clinical finding, ALS disease features, risk factors, genetics and pathophysiological mechanisms of MN degeneration. The basic scope of ALS domain ontology has been defined during interviews with neurologists involved in the OnWebDUALS research project. These meetings aimed to clarify the purpose, significance and mutual relations of the ALS patient characteristics contained in the questionnaire prepared for data collection during the OnWebDUALS project. It should be noted that the initial questionnaire content was also advised by a group of 40 expert neurologists familiar with ALS-related problems, but not involved in the project. The result of this assessment of ALS relevant notions has been described in (Mamede De Carvalho et al. 2017). It identified a consensual set of clinical data with 42 specific items that can be used as a minimal data set for patient registers and for clinical trials.

Compared to the above-mentioned minimal data set, the patient’s questionnaire ultimately applied in the OnWebDUALS project has been subsequently significantly expanded to fulfill project needs. It contains finally 156 concepts; and the corresponding questionnaire for questionnaire for control subjects contains 85 concepts. Both questionnaires are presented as the supplement files.

Final questionnaire version contains notions from areas such as: (1) demography, patient environment, life style and job history; (2) ALS clinical symptoms, dates of clinical events needed for describing disease course, selected ALS laboratory diagnostic tests, disease features, clinical phenotypes, and ALS diagnostic criteria; (3) ALS clinical signs and selected neurological manifestation not linked directly with ALS disease; (4) clinical history, including a wide range of comorbidities and clinical events which could potentially constitute ALS risk factors, such as serious traumas and surgeries, and (5) patient gene mutations in SOD1 and C9orf72 genes studied in the Project. In addition, other factors were explored, such as the pattern of disease spreading from the first affected region to other regions of the body, family history, exercise, and diagnostic track in different countries.

The core DALSO range has been defined by the final OnWebDUALS project questionnaire content and expanded by supplementing concepts existing in OnWebDUALS questionnaire and by adding new items not included in the questionnaire but necessary for the completeness and logical correctness of the modeled domain.

The final scope of DALSO is presented in details in section 3 of the paper.

### 2.3 Knowledge acquisition

The goal of the knowledge acquisition and knowledge-reuse in the ontology development process is to gather knowledge resources necessary for reliable and complete description of the model domain. Sources of knowledge can generally be classified, by reason of their form, as non-ontological and ontological. Knowledge acquisition performed during DALSO construction can be divided into three interrelated stages:

1. Establishing the possible existence of ALS domain ontology with an aim and scope at least partially corresponding to those described above; in the case of the existence of an appropriate ontology, its usefulness in the current task should be assessed, otherwise the construction of ontologies should be continued, i.e. by performing subsequent steps 2 and 3. This stage was described in section 1.2. In the absence of reusable ontological knowledge resources related to ALS, we decided to build our ontology from scratch using non-ontological knowledge and classical knowledge acquisition techniques.
2. The primary set of relevant ontology concepts has been constructed on the basis of an ALS patient questionnaire used in the OnWebDUALS Project Additional terms were collected during structured and non-structured interviews with our medical experts as well as informal or semi-formal analysis of medical texts in the field of ALS disease (risk factors—112 articles, ALS clinical knowledge—22 articles; genetic and molecular mechanisms—18 articles). All the collected terms were analyzed not only with the aim of identifying individual concepts, but also to find relationships connecting them, in order to model complex concept features and hierarchical organization of the model.
3. Searching for top-level and reference ontologies enabling knowledge reuse (5 articles).

### 2.4. Knowledge reuse – top-level and reference ontologies in DALSO

The effective implementation of knowledge reuse is based on two factors, the appropriate ontology construction i.e. normalization (described in next paper section), and application to its construction reference top-level ontologies.

Top-level and referential formal ontologies play an important role in the construction and integration of domain ontologies, providing a well-founded reference model that can be shared across domains. They form reference terminologies with a common set of abstract classes and properties that facilitate interoperability between domain ontologies and support consistent modeling decisions. To ensure knowledge reuse and interoperability with DALSO, five top-level reference ontologies have been selected.

The first upper-level ontology SCTO (El-Sappagh et al. 2018), built specifically for SNOMED-CT ontology (described below), is playing the role of general top-level reference ontology and was imported as a whole. SCTO defines the concepts of general medical science, to serve as a frame of reference for fundamental aspects of medicine such as ‘disorder’, ‘diagnosis’, ‘disease’, ‘disease course’, ‘symptom’. DALSO top-level classes are rooted in SCTO in order to avoid unclear interpretation and to enable compatibility between DALSO and other ontologies in biomedicine.

Next four biomedical reference ontologies have been used to externally reference general medical concepts in DALSO: (i) Foundational Model of Anatomy (FMA) ontology (Rosse and Mejino 2003) for human anatomy notions, (ii) Human Phenotype (HP) ontology (Robinson et al. 2008) for patient symptoms and findings, (iii) Disease Ontology (DO) (Schriml et al. 2012) for ALS patient comordibities and pathological conditions, and (iv) SNOMED-CT (Systematized Nomenclature of Medicine-Clinical Terms) ontology (Spackman 2000) was used to enhance granularity of symptoms, drugs, laboratory tests and epidemiology concepts.

The use of selected external concepts of top-level reference ontologies in DALSO meets the requirements of the minimum information to reference an external ontology term guidelines (MIREOT, Courtot et al. 2011).

## 3. DALSO general structure and information content

Medical data needed for reliable description or ALS diagnosis contain the results from many different medical knowledge areas: neurological symptoms such as upper and lower motor neuron damage, cerebellar, sensory and extrapyramidal involvement, etc., results of blood and genetic tests, electrophysiological measurements, ALS disease type classifications and clinimetric scales providing means for quantitative assessment of patient functional or mental state, and, finally, patient and his/her family medical history. These data are complemented with information relating to disease etiology and potential environmental, social and lifestyle risk factors.

To enable the construction of an ontology model, this complex and broad ALS-related medical information set should be adequately structured and modeled. The overall structure of the DALSO was based on guidelines formulated for construction of modular, normalized ontologies with explicit structure, being transparent for human readers, suitable for machine inference and supporting ontological model reuse and maintainability. Normalized ontology is an ontology organized into orthogonal modules, formed by sets of homogenous class hierarchies, joined with relations and class definitions used for restrictions and complex notions logical descriptions.

The DALSO contains 910 classes, structured in eight modules. The resulting, first level of DALSO is presented in Fig.1, together with the content of the modules and the basic relations between them. The DALSO syntactic correctness was validated by Fact++ reasoner (Tsarkov and Horrocks 2006) embedded in ontology editor applied, Protégé-2000 ver. 4.3.

**ALS Disease Description Concept** module (no. 2 in Fig. 1) is the central part of the model. To be precise, the class **ALS Disease Description** in this module can be considered as the fundamental concept in DALSO. The ALS description is expressed in categories such as disease phenotype and genotype, genetic and non-genetic risk factors and demography. Each of these general characteristics is defined as one of the remaining classes in the **ALS Disease Description Concept** module.

**Fig. 1.**
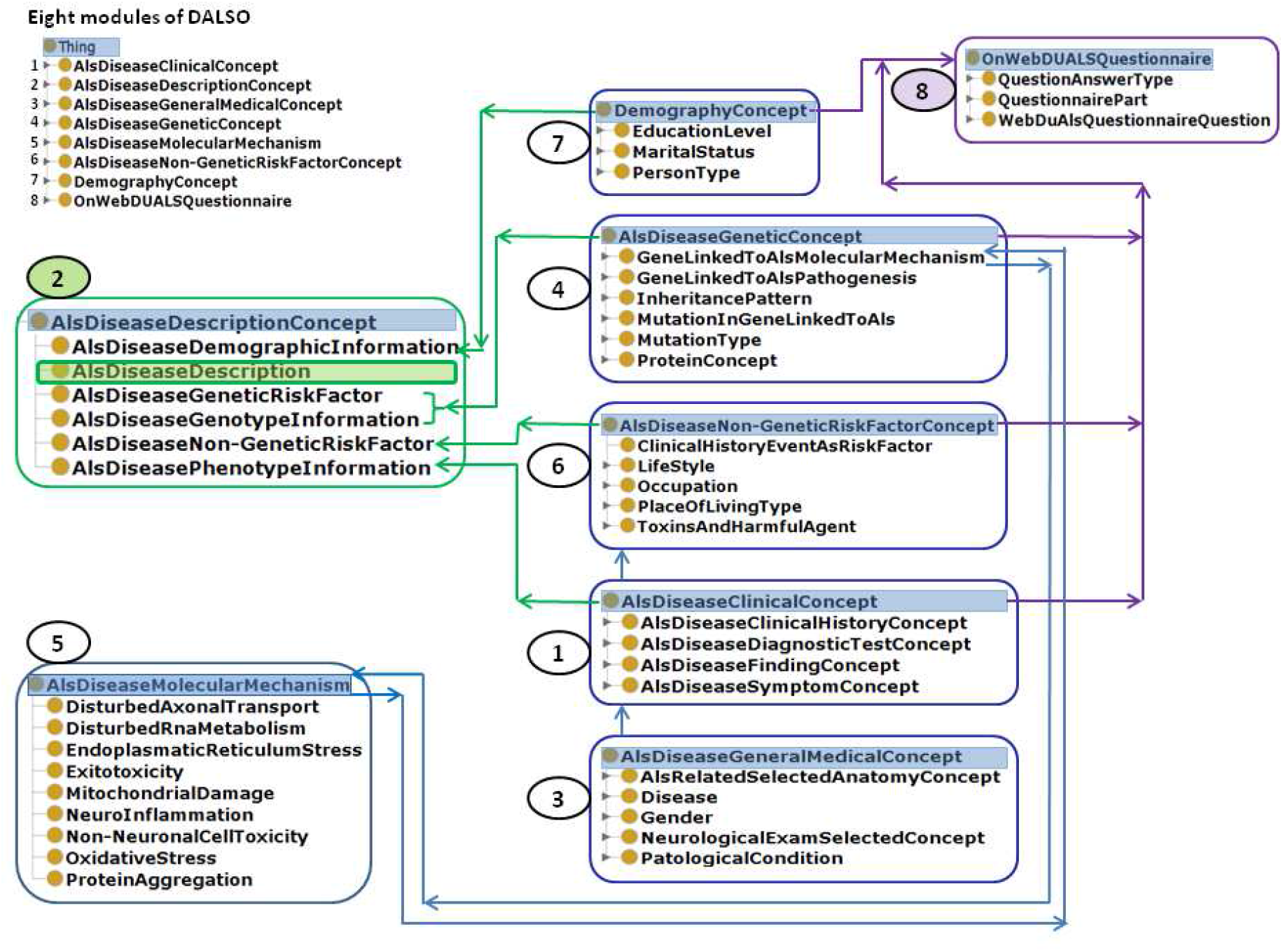
The first two levels of DALSO: modules (1^st^ level) with relations between them as shown by arrows and main classes (2^nd^ level). Left upper corner, a list of eight DALSO modules (note that all the lists are sorted alphabetically). Middle left, central DALSO module, **ALS Disease Description Concept** (2), with the class **ALS Disease Description** highlighted in green. Central part of the figure, five DALSO modules (from top to bottom: 7, 4, 6, 1, 3) providing classes defining ALS disease characteristics. Left lower corner, module of ALS Disease Molecular Mechanisms (5), right upper corner, **OnWebDUALS** Project Questionnaire modeled as ontology module 8

### 3.1 Module ALS Disease Clinical Concept

The main aim of **ALS Disease Clinical Concept** module (1) is to provide classes necessary to define the disease phenotype. These classes represent clinical history, symptoms, diagnostics tests and clinical finding. All those classes described below allow to build the definition of the class **ALS Disease Phenotype Information** in DALSO module **ALS Disease Description Concept**.

The class **ALS Disease Clinical History Concept** models important facts from the medical history of the patient, which can act as ALS risk factors encompassing comorbidities (cancer, immune disease, viral infections, cardiovascular or metabolic diseases), blood transfusion or donation, severe surgeries and traumas. All of these medical events that can be considered as ALS risk factors are used to build classes in the module **ALS Disease Non-Genetic Risk Factor Concept**.

The Class **ALS Disease Symptom Concept** contains notions defining the most common ALS patient complaints.

The class **ALS Disease Diagnostic Test Concep**t models the wide range of laboratory investigations, including blood, urine and cerebro-spinal fluid tests. Their results are used along with neurophysiology tests and neuroimaging to exclude ALS mimic syndromes, but also as ALS-specific prognostic biomarkers (Ghasemi et al. 2012; Sanderson et al. 2015; Turner et al. 2013a).

Clinical heterogeneity is a consistent feature of ALS (Al-Chalabi et al. 2016; Chiò et al. 2011; Ravits and La Spada 2009; Sabatelli et al. 2013). Major factors contributing to variable clinical phenotypes are age of onset, proportion of upper MN to lower MN deficits and unreliable prediction of disease progression (Chiò et al. 2011; Ravits and La Spada 2009; Sabatelli et al. 2013). Difficult, often delayed, diagnosis or inaccurate patient selection criteria for clinical trials, problems often encountered in the literature devoted to ALS, are partially related to ALS heterogeneity.

The content of the class **Als Disease Finding Concept** has been mainly dictated by the need of representation in DALSO the above-described ALS clinical heterogeneity. Its subclasses model concepts such as: (i) ALS clinical phenotypes based on age of onset, site of onset, disease progression rate and motor involvement; (ii) ALS diagnostic criteria, El Escorial (De Carvalho et al. 2008) and Awaji (De Carvalho and Swash 2009); (iii) ALS staging systems, Milano-Torino (MiToS) functional staging (Chiò et al. 2015) and King’s clinical staging systems (Roche et al. 2012); (iv) scales assessing ALS patient psychic and cognitive state, including ALSFRS scale (Cedarbaum et al. 1999) a validated rating tool for monitoring the progression of disability in patients with ALS; (v) ALS type (fALS, sALS) based on disease extent in patient family.

### 3.2 Module ALS Disease General Medical Concept

**ALS Disease General Medical Concept** module (3) groups five classes, used to complement the definitions of classes modeling ALS-specific clinical concepts. The class **ALS Related Selected Anatomy Concept** represents concepts necessary to define neurological clinical findings related to specific regions of the body, and concepts from clinical history. The classes **Disease** and **Pathological Condition** are used to define comordibities or events from clinical history. The class **Neurological Exam Selected Concept** gather subclasses representing concepts from the neurological examination such as motor and sensory systems, coordination, gait, and reflexes. These notions are subsequently used in the module **ALS Disease Clinical Concept (1)** to build the class **ALS Disease Finding Concept**, which finally is used to define the class **ALS Disease Phenotype Information** in the module (2).

The authors of (Gkoutos et al. 2009) stated that HP reference ontology, standardized vocabulary of phenotypic abnormalities related to human diseases, usually presents as a whole the intrinsic complexity of the defined terms. To formalize precise, fine-grained and multi-aspect knowledge implicitly inherent with clinical phenotype, it is necessary to decompose it into basic information units.

We will explain this with an example of HP ontology derived phenotype trait, characteristic for bulbar onset of ALS, *Tongue Fasciculation* (id HP:0001308), which can be broken down into the anatomical concept *Tongue Muscle* and a motor system finding *Fasciculation*. Here, the elementary features are: (i) localization in two anatomic structures, tongue muscle and head; (ii) localization in bulbar region, an anatomic area specific for ALS; (iii) the relation of the modeled phenotypic trait to specific MN type, in this case the lower MN. They have been modeled as subclasses of the root class **ALS Disease Selected Anatomy Concept**. The relation of the class Tongue Muscle Fasciculation with its features is modeled via four object type properties. The resulting *Tongue Fasciculation* composite concept is defined at a higher level of detail, allowing for more precise phenotype description. The definition of the class Tongue Muscle Fasciculation is presented in Fig. 2.

**Fig. 2.**
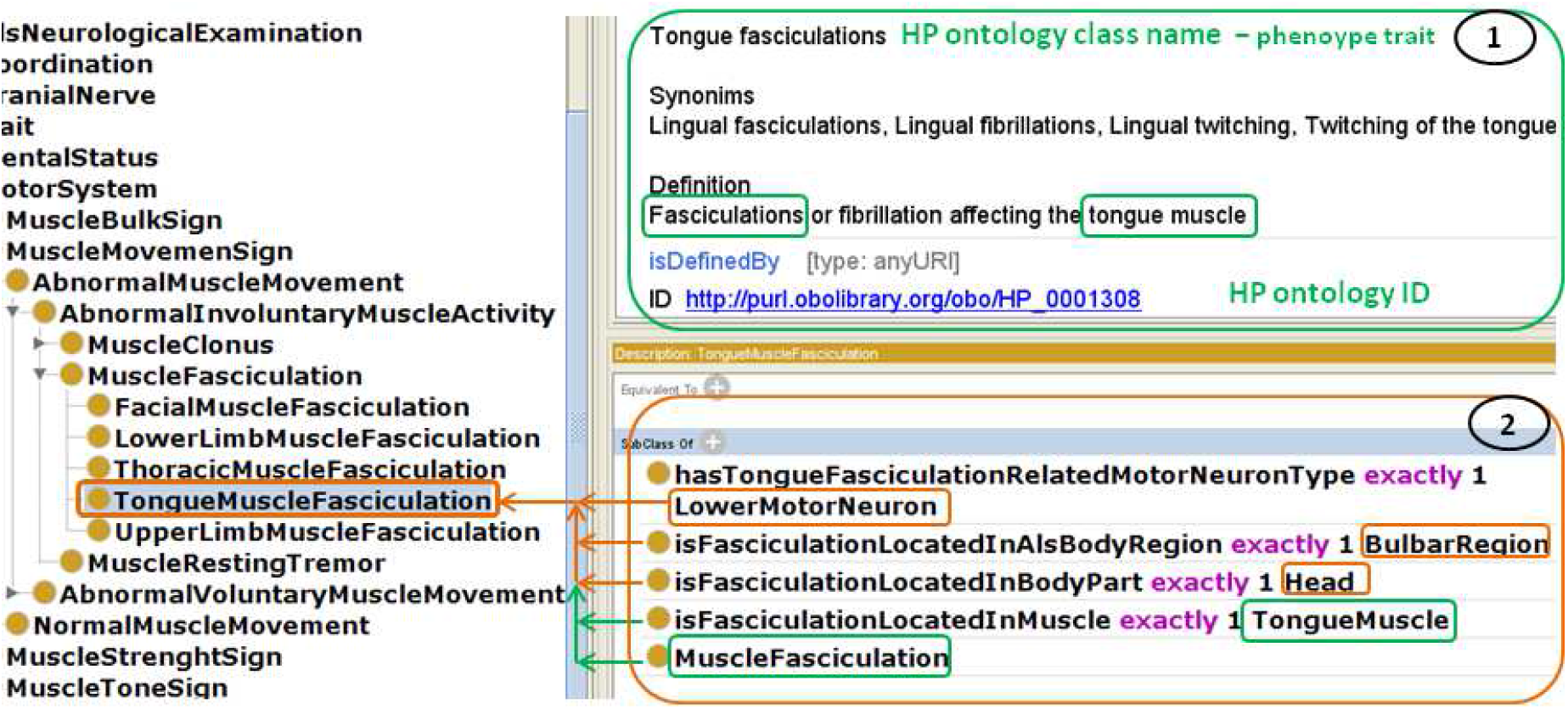
The definition of the class Tongue Muscle Fasciculation in DALSO (screenshot from ontology editor Protege-2000). On the left, a fragment of class hierarchy, defined class highlighted in orange. Green panel (1), references from HP ontology to the class Tongue Muscle Fasciculation: HP term for a phenotype trait, HP identifier and textual definition. Orange panel (2), details of the class Tongue Muscle Fasciculation definition. Notions implicitly present in HP definition, Tongue Muscle and Fasciculation (marked with green) are made explicit, additional notions *Lower Motor Neuron, Bulbar Region* and *Head* are also made explicit (marked with orange). The resulting definition of *Tongue Fasciculation* in DALSO allows for deeper understanding of modeled phenotype trait.

### 3.3 Module ALS Disease Non-Genetic Risk Factor

Although many risk factors have been proposed and studied for ALS, little progress has been made in identifying those with a high degree of causality (Wang et al. 2017). Current research suggests that the only established risk factors for ALS are family history of ALS (fALS), male gender and older age. Identification of non-genetic ALS risk factors, i.e. environmental, toxic, infectious or metabolic etiologies of ALS, is highly relevant for detecting mechanisms of susceptibility to the sporadic form of ALS.

The knowledge regarding risk factors has been collected as a result of discussions among OnWebDUALS experts and a review on the literature on ALS risk factors was prepared in frames of the ONWebDUALS Project (eventually published as an opinion article, Kuraszkiewicz et al. 2018). Most of the authors point to possible ALS non-genetic risk factors that can be divided into the following four main groups: related to lifestyle, occupation, environment or medical history. However, the research in this area is generally inconclusive.

The class **LifeStyle** represents lifestyle-related ALS risk factors, including smoking (Armon 2003; Das et al. 2012; Turner 2013), dietary factors, nutritional status, vitamin deficiency (Rosenfeld and Ellis 2008; Verburgh 2014), physical fitness and body-mass index (Armon 2003; Turner 2013).

The class **Occupation** models in DALSO occupational risk-factors, as e.g., exposure of industry and construction workers to heavy metals, electromagnetic fields and toxic chemicals, such as solvents and dyes or exposure of soldiers during military service to chemicals used in battlefields, severe injuries and stress. The class **Place Of Living** contains environmental risk factors, such as residential exposure to electromagnetic fields or to chemicals, including pesticides (Beard et al. 2016; Cragg et al. 2017; Das et al. 2012; Park et al. 2005; Su et al. 2016; Tai et al. 2017; Weisskopf et al. 2009).

The class **Clinical History Event As Risk Factor** represents selected medical history facts that can act as ALS risk factors, including comorbidities, e.g. viral infections (Alfahad and Nath 2013; Oluwole et al. 2007), autoimmune diseases (Turner et al. 2013a) or cancer (Freedman et al. 2013; Gibson et al. 2016), medical conditions such as leaky intestine (De Marchi et al. 2018; Kuraszkiewicz et al. 2018; Rowin et al. 2017; Wright et al. 2018), head and spine traumas or surgeries, electrical injuries (Berger et al. 2000; Chen et al. 2007; Das et al. 2012; Turner et al. 2013b) or stress (Okamoto et al. 2009).

Four above described classes representing ALS non-genetic risk factor serve in turn to build the definition of the class **ALS Disease Phenotype Information** in the module (2). Class **Toxins And Harmful Agent** is an auxiliary class that models concepts such as chemical, physical or emotional stressors, used to complement classes representing four main ALS disease risk factors.

### 3.4 Module ALS Disease Genetic Concept

The module **ALS Disease Genetic Concept (4)** gathers classes representing genetic concepts included in DALSO. The class **Gene Linked To ALS Pathogenesis** groups as subclasses genes related to ALS pathogenesis, listed in Amyotropic Lateral Sclerosis Online Genetics Database (ALSoD, http://alsod.iop.kcl.ac.uk/) (Abel et al. 2012). The authors of (Fundel and Zimmer 2006) revealed the existence of significant ambiguity within gene names and showed that the degree of overlap between different public data sources is generally relatively modest.

Thus, to ensure explicitness in gene identity, all subclasses of the class **Gene Linked To ALS Pathogenesis** have three datatype properties ensuring their unambiguous identification. These are: (1) gene name from ALSoD, (2) HUGO Nomenclature Committee (HGNC) (Wain et al. 2002) identifier and (3) identifier from the online catalogue of human genes and genetic disorders (Online Mendelian Inheritance in Man, OMIM, Hamosh et al. 2002), https://www.ncbi.nlm.nih.gov/omim). Fourth data type property of the class representing genes involved in ALS pathogenesis indicates the gene location within the chromosome. The class definition is completed by relations with two important model concepts, the classes **Mutations In Gene Linked To ALS** and **Protein Concept**.

The class **Mutation In Gene Linked To ALS** gathers sub-classes representing mutations in genes related to ALS pathogenesis. Their definitions are based on the guidelines of the Mutation Nomenclature of the Human Genome Variation Society (den Dunnen and Antonarakis 2000) and also on dependencies defined in the DALSO. According to mutation nomenclature, the gene mutation is defined using four datatype properties representing mutation, its original and mutated content, offset and count. Type of the mutation is modeled as the object type relation linking class representing mutation with the class **Mutation Type**. The association of the mutation and the gene is represented via the object type relation between the classes representing gene mutation and gene.

The class **Protein Concept** is defined by its name, function and relation to the gene by which the protein is encoded. It is linked with the module **ALS Disease Molecular Mechanism** (5).

### 3.5 Module ALS Disease Description Concept

As it was stated in section 3.1, **ALS Disease Description Concept** module with its class **ALS Disease Description** constitutes the most important part of the DALSO. The class **ALS Disease Description** is characterized along five axes, phenotype and genotype information, genetic and non-genetic risk factors and demography (Fig. 3). Each axis is represented as one class in the module, with a class name indicating modeled content (see Fig.1). Each class modeling specified axis relates to a specific ontology module, gathering classes modeling its structure and content.

**Fig. 3.**
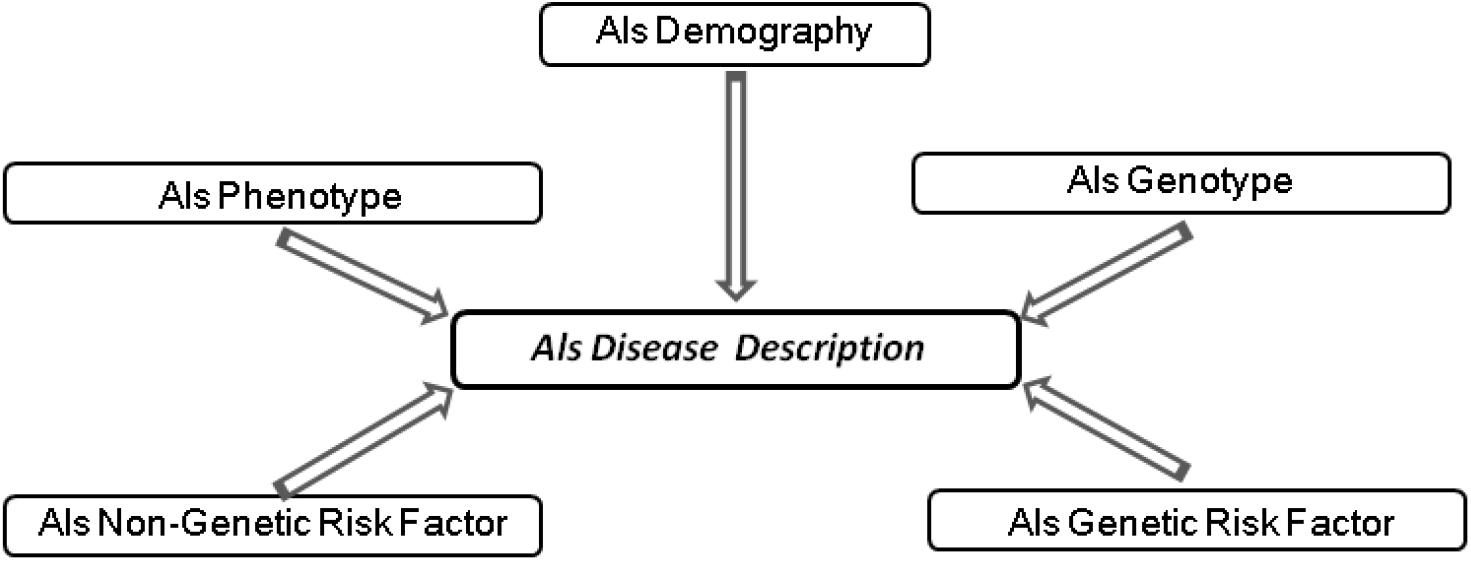
General overview of the class **ALS Disease Description** structure. ALS is characterized along five information axes, phenotype and genotype information, genetic and non-genetic risk factors and demography.

### 3.6 Module Als Disease Molecular Mechanism

The module **ALS Disease Molecular Mechanism** (5) contains classes modeling mechanisms involved in MN degeneration. As many works indicate (Ferraiuolo et al. 2011; Shaw 2005; Taylor et al. 2016; Morgan and Orrell 2016; S. Chen et al. 2013; Souza et al. 2015; Pasinelli and Brown 2006; Zhou et al. 2017), mechanisms underlying MN death are multifactorial and MN degeneration probably results from a complex interplay of multiple mechanisms. The analysis of the cited above literature allowed distinguishing nine basic mechanisms involved in the pathway leading to the MN death:

1. Oxidative stress,
2. Excitotoxicity,
3. Mitochondrial damage,
4. Neuroinflammation,
5. Disturbed axonal transport,
6. Endoplasmic reticulum stress,
7. Protein aggregation,
8. Disturbances in RNA metabolism,
9. Non-neuronal cell dysfunction.

The complete picture of the pathogenic mechanisms of ALS is still unclear, thus true participation and significance of the above-listed putative mechanisms may in fact effectively vary in particular patients or within their subgroups. However, a deep understanding of the complex interplay between multiple processes is critical for development of new therapeutic approaches. This consideration motivated us to include in DALSO the module representing MN degeneration pathways. Many recent studies state that dysfunctions of vital molecular pathways underlying ALS pathogenesis are closely related to genetic factors, namely to mutations in specific genes (Pasinelli and Brown 2006; Chen et al. 2013; Souza et al. 2015; Morgan and Orrell 2016; Taylor et al. 2016). Each class in the module **ALS Disease Molecular Mechanism** is related to the class **Gene Linked to ALS Molecular Mechanism** in the module **(4)** representing specific genes involved in specific mechanism of motor neuron degeneration.

### 3.7 Modules OnWebDUALS Questionnaire and Demography Concept

The module **OnWebDUALS Questionnaire (8)** represents the content of questionnaire items used in the OnWebDUALS project (see supplement file), defined using ontology concepts, except for concepts modeling the time periods and temporal sequence of events.

The module **Demography Concept (7)**, the smallest DALSO module, gathers rudimentary demographic concepts, education level, marital status and family members. Its classes are used to model concepts in OnWebDUALS questionnaire and demographic information in the **ALS Disease Description** module (**2**, see Fig.1).

## 4. Conclusions

The DALSO is so far the only model representing a comprehensive, formal, structured knowledge source for the ALS domain. The ontology includes a wide range of ALS-related aspects, ranging from clinical information and clinical history data, to disease mechanisms and genetics. DALSO is the first ALS disease ontological model and it complements the spectrum of existing domain disease ontologies referring to neurodegenerative diseases (Malhotra et al. 2014; Younesi et al. 2015). The model shortcoming is a lack of concepts that allow modeling of the temporal sequence of events.

The DALSO is expressed in OWL2 language, contains 910 classes, is consistent and free of logical errors. Its syntactic correctness was validated by the Fact++ reasoner (Tsarkov and Horrocks 2006) embedded in Protégé-2000 ver 4.3 ontology editor, used to develop the model.

The DALSO provides the means for defining schemes of ALS patient resources and also for defining schemes for clinical trials. DALSO ontology used in combination with Ontop, an Ontology Based Data Access system (Calvanese et al. 2017a), can be used to access a relational database through ontological conceptual layers. The efficiency of this approach was proven by Zhang et al. (2018) for the integration of cancer data. DALSO can also be used for creation of a practicable health informatics service system (Djedidi and Aufaure 2007; Silachan and Tantatsanawong 2011).

Another potential application of DALSO is in refined versions of enrichment analysis, proposed by (Shah et al. 2012), who suggests the next step of genetic data analysis could be e.g. replacement of GO ontology with specific disease ontology.

Summing up, the DALSO can be used as a starting point for the development of systems for the diagnosis support, education of young doctors in this field, and for the integration and analysis of clinical, genetic and other data collected in various databases (Ivanović and Budimac 2014). This could help speed up the research on the ALS etiology and development of effective treatments for this lethal disease.

The final version of DALSO was created using the Protégé-2000 ver 4.3 editor. It can be uploaded at the link https://als.ibib.waw.pl/DAlsO.zip. Use of the DALSO ontology is free of charge, provided that any future publication based on its use cites this article.

## Data Availability

All data produced are availale at https://als.ibib.waw.pl/DAlsO.zip

## Acknowledgment

This paper was supported by the grant JPND/01/2015 funded by the Polish National Center of Research and Development in frames of EU Joint Program of Neurodegenerative Research (JPND).

