## Supplemental tables for "DALSO: domain ALS ontology": Questionnaire_Control.pdf (from another version of this preprint: version 1, 2024-06-03)

| Nº | 1 – GENERAL DATA | | | | |
|---|---|---|---|---|---|
| 1 | **ONWebDUALS site** | | | | |
| 2 | **Physician recording the data** | | | | |
| 3 | **Patient code name** | | | | |
| 4 | **Date of Consultation** (day-month-yy) | | | | |
| | **2 – TIMES** | | **Not feasible** | **Not available** | **Not relevant** |
| 5 | **Date of birth** (day-month-yy) | | | | |
| 6 | **Gender** (F female, M male) | F / M | | | |
| 7 | **Ethnicity**<br>C Caucasian; AF African-origin, AS Asian | C / AF / AS | | | |
| 8 | **Place of Birth** (city / country)<br>(One choice) | ………………………/……………………..<br>Village - < 1000 inhabitants<br>Country towns– 1000-5 000<br>Small town– 5 000-20 000<br>Middle town– 20 000 – 100 000<br>Large town– > 100 000 | | | |
| 8’ | **Place of Birth** characteristics (one choice) | Rural area vs urban area<br>(an urban area is the region surrounding a city). | | | |
| 9 | **Mother´s Place of Birth** (city / country) | ………………………/……………………..<br>Village - < 1000 inhabitants<br>Country towns– 1000-5 000<br>Small town– 5 000-20 000<br>Middle town– 20 000 – 100 000<br>Large town– > 100 000 | | | |
| 9’ | **Place of Birth** characteristics (one choice) | Rural area vs urban area | | | |
| 10 | **Father´s Place of Birth** (city / country) | ………………………/……………………..<br>Village - < 1000 inhabitants<br>Country towns– 1000-5 000<br>Small town– 5 000-20 000<br>Middle town– 20 000 – 100 000<br>Large town– > 100 000 | | | |
| 10’ | **Place of Birth** characteristics (one choice) | Rural area vs urban area | | | |
| | **3 – CO-MORBIDITIES (past and present)** | | Not feasible | Not available | Not relevant |
| 11 | **Blood hypertension** | Yes / No | | | |
| 12 | **Diabetes – type I / II** | Yes (type 1 vs type 2) / No | | | |
| 13 | **Hypercholesterolemia** | Yes / No | | | |
| 14 | **Hypertriglyceridemia** | Yes / No | | | |
| 15 | **Hyperthyroidism** | Yes / No | | | |
| 16 | **Hypothyroidism** | Yes / No | | | |
| 17 | **Autoimmune rheumatologic disorder** | Yes / No | | | |
| 18 | **Autoimmune intestinal disorder** | Yes / No | | | |
| 19 | **Stroke** | Yes (ischemic vs hemorrhagic vs unknown) / No | | | |
| 20 | **Heart** – ischemia | Yes / No | | | |
| 21 | **Heart** – arrhytmia | Yes / No | | | |
| 22 | **Heart** – insufficiency | Yes / No | | | |

| | | | | | |
|---|---|---|---|---|---|
| 23 | **Cancer (primary)**<br>(multiple choice) | No / Yes:<br>Brain (B); Neck (N); Breast (Br); Lung (L); other Thoracic (T); Gastric (G); Colon (C); Rectum (Re); other Abdominal (A); Prostate (Pr); Uterus (U); other Pelvic (P); Spine (Sp); Bone cancer and sarcomas in upper limbs (UL); Bone cancer and sarcomas in lower limbs (LL); Skin (S); Leukemia (L); Lymphoma (Ly); other blood cancer (Bl). | | | |
| 23' | **Metastasis** (above list) | | | | |
| 24 | **Cancer treatment – Chemotherapy**<br>Specify the drug if possible | Yes / No<br>…………………………………………… | | | |
| 25 | **Cancer treatment – Radiotherapy**<br>**Head or neck** | Yes / No<br>Yes / No | | | |
| 26 | **Smoking**<br>Date of initiation (year)<br>Stopped<br>Date of interruption (year)<br>Mean number of packs/day | Yes / No<br>……………………………………………<br>Yes / No<br>……………………………………………<br>…………………………………………… | | | |
| | **4 – MEDICATION** (generic name) | | Not feasible | Not available | Not relevant |
| 27 | **Psychiatric** (> 6 months anytime)<br>Specify the drug if possible | Yes / No<br>…………………………………………… | | | |
| 28 | **Supplements** (> 6 months anytime)<br>Specify if possible | Yes / No<br>…………………………………………… | | | |
| 29 | **Riluzole**<br>Date of initiation<br>Side effects? | Yes / No<br>……………………………………………<br>…………………………………………… | | | |
| 30 | **Statins** (> 6 months anytime)<br>Specify if possible | Yes / No<br>…………………………………………… | | | |
| 31 | **Antiepileptic drug** (> 6 months anytime)<br>Specify if possible | Yes / No<br>…………………………………………… | | | |
| 32 | **NSAID** (> 6 months anytime)<br>Specify if possible | Yes / No<br>…………………………………………… | | | |
| 33 | **Steroids** (> 6 months anytime)<br>Specify if possible | Yes / No<br>…………………………………………… | | | |
| 34 | **Immunosupressors** (> 6 months anytime)<br>Specify the drug if possible | Yes / No<br>…………………………………………… | | | |
| | **5 – GENETIC INFORMATION** | | Not feasible | Not available | Not relevant |
| 35 | **SOD1 mutation** (specify) | Yes…………………………………… / No | | | |
| 36 | **Abnormal C9orf72 repeat-expansion** | Yes / No | | | |
| 37 | **Other genes**<br>**Was the patient included in genomic studies** (specify) | Yes/ No<br>Yes…………………………………… / No | | | |
| 38 | **Familial history of ALS** (Multiple choice) | No<br>mother<br>father<br>heterozygous twin brother<br>homozygous twin brother<br>heterozygous twin sister<br>homozygous twin sister<br>Other family members:<br>…………………………………………………………………<br>…………………………………………………………………<br>…………………………………………………………………<br>…………………………………………………………………<br>…………………………………………………………………<br>………………………………………………………………… | | | |

| | | |
|---|---|---|
| 39 | **Familial history of FTD** | No<br>mother<br>father<br>heterozygous twin brother<br>homozygous twin brother<br>heterozygous twin sister<br>homozygous twin sister<br>Other family members:<br>……………………………………………………………………<br>……………………………………………………………………<br>……………………………………………………………………<br>……………………………………………………………………<br>……………………………………………………………………<br>…………………………………………………………………… |
| 40 | **Familial history of Alzheimer disease** | No<br>mother<br>father<br>heterozygous twin brother<br>homozygous twin brother<br>heterozygous twin sister<br>homozygous twin sister<br>Other family members:<br>……………………………………………………………………<br>……………………………………………………………………<br>……………………………………………………………………<br>……………………………………………………………………<br>……………………………………………………………………<br>…………………………………………………………………… |
| 41 | **Familial history of Parkinson disease** (which family member) | No<br>mother<br>father<br>heterozygous twin brother<br>homozygous twin brother<br>heterozygous twin sister<br>homozygous twin sister<br>Other family members:<br>……………………………………………………………………<br>……………………………………………………………………<br>……………………………………………………………………<br>……………………………………………………………………<br>……………………………………………………………………<br>…………………………………………………………………… |
| 42 | **Familial history of multiple sclerosis** (which family member) | No<br>mother<br>father<br>heterozygous twin brother<br>homozygous twin brother<br>heterozygous twin sister<br>homozygous twin sister<br>Other family members:<br>……………………………………………………………………<br>……………………………………………………………………<br>……………………………………………………………………<br>……………………………………………………………………<br>……………………………………………………………………<br>…………………………………………………………………… |
| 43 | **Other severe diseases in the family** | 1. Mother……………………………………………………<br>……………………………………………………………………<br>……………………………………………………………………<br>……………………………………………………………………<br>2. Father……………………………………………………<br>…………………………………………………………………… |

| | | | | | | |
|---|---|---|---|---|---|---|
| | | ……………………………………………………………<br>3. ………………………………………………………<br>4. ………………………………………………………<br>5. ………………………………………………………<br>6. ………………………………………………………<br>7. ………………………………………………………<br>8. ………………………………………………………<br>9. ………………………………………………………<br>10.……………………………………………………… | | | | |
| 44 | **Parents' and siblings´ causes of death** | 1. Mother: Alive Yes / No<br>……………………………………………<br>2. Father: Alive Yes / No<br>……………………………………………<br>3. ……………………………………………<br>4. ……………………………………………<br>5. ……………………………………………<br>6. ……………………………………………<br>7. ……………………………………………<br>8. ……………………………………………<br>9. ……………………………………………<br>10. ………………………………………… | Age<br>………<br><br>………<br>………<br>………<br>………<br>………<br>………<br>………<br>………<br>……… | | | |
| 45 | **Mother´s number of siblings and birth order** (e.g. 2nd child in 7, should be 2/7) | | | | | |
| 46 | **Father´s number of siblings and birth order** (e.g. 2nd child in 7, should be 2/7) | | | | | |
| 47 | **Consanguinity of parents** | Yes / No | | | | |
| 48 | **Number of Siblings and birth order** (e.g. 2nd child in 7, should be 2/7)<br>With monozygous twin<br>With a dizygous twin | ……………………………………………<br>Yes / No<br>Yes / No | | | | |
| | **6 – HABITS/TRAUMA AND SURGERY** | | | **Not feasible** | **Not available** | **Not relevant** |
| 49 | **Regular physical exercise** | Yes / No | | | | |
| 50 | **Intense physical exercise** > 1 year<br><br>1: 150 min/week of moderate aerobic activity (such as fast walking, swimming) | **Which sport:**<br>Basketball (BA); Boxing(BO); Climbing (CL); Cycling (CY); Dancing (D); Diving (DV); Fast walking (FW); Fencing (FE); Fitness (F); Football (FB); Golf (G); Handball (HB); Hockey (HO); Rowing (RO); Rugby (RU); Running (R); Skiing (SK); Swimming (S); Tennis (T); Volleyball (V); Weightlifting(WL)<br>Others: | **How long (yrs):**<br>e.g.: DV: 10, SK 25 (diving 10 yrs, skiing 25 yrs) | | | |
| | 2: 75 min/week of vigorous aerobic activity (such as running or aerobic dancing) | **Which sport:**<br>Basketball (BA); Boxing(BO); Climbing (CL); Cycling (CY); Dancing (D); Diving (DV); Fast walking (FW); Fencing (FE); Fitness (F); Football (FB); Golf (G); Handball (HB); Hockey (HO); Rowing (RO); Rugby (RU); Running (R); Skiing (SK); Swimming (S); Tennis (T); Volleyball (V); Weightlifting(WL)<br>Others: | **How long (yrs):** | | | |

| | | | |
|---|---|---|---|
| 51 | **Mild physical exercise** > 1 year<br>(< intensity above) | **Which sport:**<br>Basketball (BA); Boxing(BO); Climbing (CL); Cycling (CY); Dancing (D); Diving (DV); Fast walking (FW); Fencing (FE); Fitness (F); Football (FB); Golf (G); Handball (HB); Hockey (HO); Rowing (RO); Rugby (RU); Running (R); Skiing (SK); Swimming (S); Tennis (T); Volleyball (V); Weightlifting(WL)<br>Others: | **How long (yrs):** |
| 52 | **Regular specific diet** > 1 year (specify) | No<br>Mediterranean (M); Vegetarian (VT); Vegan (VG); Gluten-free (GF); High protein (HP); Low carbohydrate (LC); Paleo (High- protein, high-fat low-carbohydrate) (PA) | |
| 53 | **Head/Neck trauma** in the last 5 years before data entry (only if hospitalized) | Yes (head vs neck) / No | |
| 54 | **Head/Neck trauma** > 5 years before data entry (only if hospitalized) | Yes (head vs neck) / No | |
| 55 | **Other trauma** in the last 5 years (only if hospitalized)<br>Cervical (C)<br>Thoracic (T)<br>Lumbo-sacral (LS) | Yes (C, T, L-S) / No | |
| 56 | **Other trauma** > 5 years before data entry (only if hospitalized)<br>Cervical (C)<br>Thoracic (T)<br>Lumbo-sacral (LS) | Yes (C, T, L-S) / No | |
| 57 | **Spine surgery** in the last 5 years before data entry<br>Cervical (C)<br>Thoracic (T)<br>Lumbo-sacral (LS) | Yes (C, T, L-S) / No | |
| 58 | **Spine surgery** > 5 years before data entry<br>Cervical (C)<br>Thoracic (T)<br>Lumbo-sacral (LS) | Yes (C, T, L-S) / No | |
| 59 | **Limb surgery**<br>in the last 5 years before data entry<br>Upper limb (UL)<br>Lower lib (LL) | Yes (UL, LL) / No | |
| 60 | **Limb surgery** > 5 years before data entry<br>Upper limb (UL)<br>Lower limb (LL) | Yes (UL, LL) / No | |
| 61 | **Abdominal surgery** in the last 5 years before data entry | Yes / No | |
| 62 | **Abdominal surgery**<br>> 5 years before data entry | Yes / No | |
| 63 | **Thoracic surgery**<br>in the last 5 years before data entry | Yes / No | |
| 64 | **Thoracic surgery**<br>> 5 years before data entry | Yes / No | |
| 65 | **Pelvic Surgery**<br>in the last 5 years before data entry | Yes / No | |
| 66 | **Pelvic Surgery**<br>> 5 years before data entry | Yes / No | |

| | | | | | |
|---|---|---|---|---|---|
| 67 | **Head/neck Surgery**<br>in the last 5 years before data entry | Yes / No | | | |
| 68 | **Head/neck Surgery**<br>> 5 years before data entry | Yes / No | | | |
| 69 | **Blood transfusion**<br>in the last 5 years before data entry | Yes / No | | | |
| 70 | **Blood transfusion**<br>> 5 years before data entry | Yes / No | | | |
| 71 | **Blood donor**<br>in the last 5 years before data entry | Yes / No | | | |
| 72 | **Blood donor**<br>> 5 years before data entry | Yes / No | | | |
| 73 | **Electric shock** in the last 5 years before data entry (only if hospitalized) | Yes / No | | | |
| 74 | **Electric shock** > 5 years before data entry (leading to hospitalization) | Yes / No | | | |
| 75 | **Emotional stress** in the last 5 years before data entry (loss or disease of close family member, loss of job ) | Yes / No | | | |
| 76 | **Emotional stress** >5 years before data entry (loss or disease of close family member, loss of job ) | Yes / No | | | |
| | **7 – OCCUPATIONS** | | **Not feasible** | **Not available** | **Not relevant** |
| 77 | **Main Occupation**<br>in the last 5 years before data entry | | | | |
| 78 | **Main Occupation**<br>>5 years before data entry | | | | |
| 79 | **Second Occupation**<br>in the last 5 years before data entry | | | | |
| 80 | **Second Occupation**<br>> 5 years before data entry | | | | |
| 81 | **Place of living**<br>in the last 5 years before data entry | ............................/..........................<br>Village - < 1000 inhabitants<br>Country towns– 1000-5 000<br>Small town– 5 000-20 000<br>Middle town– 20 000 – 100 000<br>Large town– > 100 000 | | | |
| 81’ | **Place of living characteristics**<br>in the last 5 years before data entry<br>(multiple choice) | Rural area (RA); Urban area (UA);<br>Near sewage plant (NSP);<br>Near landfills (NL);<br>Near waste incinerators (MWI);<br>Near sources of EM fields (high-voltage transmission lines, radar etc.) (EMF) | | | |
| 82 | **Place of living**<br>> 5 years before data entry | ............................/..........................<br>Village - < 1000 inhabitants<br>Country towns– 1000-5 000<br>Small town– 5 000-20 000<br>Middle town– 20 000 – 100 000<br>Large town– > 100 000 | | | |
| 82’ | **Place of living characteristics**<br>> 5 years before data entry<br>(multiple choice) | Rural area (RA); Urban area (UA);<br>Near sewage plant (NSP);<br>Near landfills (NL);<br>Near waste incinerators (MWI);<br>Near sources of EM fields (high-voltage transmission lines, radar etc.) (EMF) | | | |
| 83 | **ALS cases in the neighborhood** | Yes / No | | | |
| 84 | **ALS disease in co-workers** | Yes / No | | | |
| 85 | **ALS disease in friends** | Yes / No | | | |
