## Supplemental tables for "DALSO: domain ALS ontology": Questionnaire_Patient.pdf (from another version of this preprint: version 1, 2024-06-03)

| Nº | 1 – GENERAL DATA | | | | |
|---|---|---|---|---|---|
| 1 | **ONWebDUALS site** | | | | |
| 2 | **Physician recording the data** | | | | |
| 3 | **Patient code name** | | | | |
| 4 | **Date of Consultation** (day-month-yy) | | | | |
| | **2 – TIMES** | | **Not feasible** | **Not available** | **Not relevant** |
| 5 | **Date of birth** (day-month-yy) | | | | |
| 6 | **Gender** (F female, M male) | F / M | | | |
| 7 | **Ethnicity**<br>C Caucasian; AF African-origin, AS Asian | C / AF / AS | | | |
| 8 | **Place of Birth** (city / country)<br>(One choice) | ……………………../……………………..<br>Village - < 1000 inhabitants<br>Country towns– 1000-5 000<br>Small town– 5 000-20 000<br>Middle town– 20 000 – 100 000<br>Large town– > 100 000 | | | |
| 8' | **Place of Birth** characteristics (one choice) | Rural area vs urban area<br>(an urban area is the region surrounding a city). | | | |
| 9 | **Mother´s Place of Birth** (city / country) | ……………………../……………………..<br>Village - < 1000 inhabitants<br>Country towns– 1000-5 000<br>Small town– 5 000-20 000<br>Middle town– 20 000 – 100 000<br>Large town– > 100 000 | | | |
| 9' | **Place of Birth** characteristics (one choice) | Rural area vs urban area | | | |
| 10 | **Father´s Place of Birth** (city / country) | ……………………../……………………..<br>Village - < 1000 inhabitants<br>Country towns– 1000-5 000<br>Small town– 5 000-20 000<br>Middle town– 20 000 – 100 000<br>Large town– > 100 000 | | | |
| 10' | **Place of Birth** characteristics (one choice) | Rural area vs urban area | | | |
| 11 | **Date of 1st Symptoms** (month-yy)<br>1st symptoms reports to weakness or cognitive changes | | | | |
| 12 | **Date of 1st Medical Consultation related to the ALS symptoms** (day-month-yy) | | | | |
| 13 | **Date of Diagnosis** (day-month-yy)<br>Patient informed about the disease | | | | |
| 14 | **Date of Invasive ventilation** or **Non-Invasive ventilation ≥ 23hours/day** (day-month-yy) | | | | |
| 15 | **Date of Death** (day-month-yy) | | | | |
| | **3 – DISEASE FEATURES** | | **Not feasible** | **Not available** | **Not relevant** |
| 16 | **Region of onset** – multiple choice<br>first symptoms reports to weakness | Limbs Bulbar Neck<br>Thoracic / Abdominal<br>Respiratory/ Generalized | | | |
| 17 | **Predominant UMN vs LMN manifestation at onset** | UMN / LMN | | | |
| 18 | **Limb onset** | No<br>upper limb<br>lower limb \| **Predominant side:**<br>left, right, symmetric \| **Predominant impairment:**<br>distal<br>proximal | | | |

| | | | | | |
|---|---|---|---|---|---|
| 19 | **Fasciculations at onset** | Yes / No | | | |
| 20 | **Weight loss (>10% initial weight)**<br>Yes / No<br>If yes, when (one choice) | < 6 months before disease onset<br>6-12 months before disease onset<br>12-24 months before disease onset<br>24-36 months before disease onset | | | |
| 21 | **Emotional lability at onset**<br>(symptom or clinical sign) | Yes / No | | | |
| 22 | **Cognitive symptoms at onset**<br>Yes / No<br>If yes, which (multiple choice) | behavioral symptoms<br>language deficit<br>other manifestations | | | |
| 23 | **Handedness** (left (L) vs right (R) vs ambidextrous (A)) | L / R / A | | | |
| | **4 – CLINICAL SIGNS** (at study entry) | | **Not feasible** | **Not available** | **Not relevant** |
| | **4.1 Bulbar region** | | | | |
| 24 | Bulbar UMN sign (multiple choice) | tongue spasticity<br>jaw clonus<br>brisk jaw jerk | | | |
| 25 | Bulbar LMN sign (multiple choice) | tongue atrophy<br>tongue fasciculations<br>weak orbicularis oris<br>facial muscle fasciculations<br>masseter atrophy | | | |
| | **4.2 Upper limbs** | | | | |
| 26 | UMN sign - | Hyperreflexia<br>Finger flexion / Hoffman sign<br>Spasticity | | | |
| 27 | LMN sign - | Atrophy and weakness<br>Fasciculations<br>Hyporeflexia | | | |
| | **4.3 Lower limbs** | | | | |
| 28 | UMN sign (multiple choice) | Hyperreflexia<br>Babinski's sign<br>Spasticity | | | |
| 29 | LMN sign (multiple choice) | Atrophy and weakness<br>Fasciculations<br>Hyporeflexia | | | |
| | **4.4 Axial Muscles** | | | | |
| 30 | Neck weakness | Yes / No | | | |
| 31 | Thoracic muscle fasciculations | Yes / No | | | |
| 32 | Resting respiratory fatigue or Exertional respiratory fatigue to minor efforts | Yes / No | | | |
| 33 | Orthopnea | Yes / No | | | |
| 34 | Paradoxical respiration | Yes / No | | | |
| 35 | Weak cough | Yes / No | | | |
| | **4.5 Diagnosis Category** | | | | |
| 36 | Revised El Escorial Criteria (single choice) | Definite; Probable; Possible;<br>Probable-laboratory supported | | | |
| 36’ | Progressive muscle atrophy (PMA)<br>Primary lateral sclerosis (PLS)<br>Monomelic (M)<br>Progressive Bulbar Palsy (PBP) | Yes / No<br>Yes / No<br>Yes / No<br>Yes / No | | | |

| | **4.6 Other Neurological Manifestations** | | | | | | |
|---|---|---|---|---|---|---|---|
| 37 | Emotional lability | Yes / No | | | | | |
| 38 | Extrapyramidal signs (resting tremor) in | Upper Limbs / Lower Limbs / No | | | | | |
| 38´ | Extrapyramidal signs (bradykinesia) in | Upper Limbs / Lower Limbs / No | | | | | |
| 39 | Sensory changes in<br>(objective changes on neurological testing) | Upper Limbs / Lower Limbs / No | | | | | |
| 40 | Apraxia (facial) | Yes / No | | | | | |
| 41 | Apraxia (upper limbs) | Yes / No | | | | | |
| 42 | Postural instability –<br>Nutt Unexpected Retropulsive Test<br>(if independent walk is preserved) | Positive / Negative | | | | | |
| 43 | Broad-based gait (if legs are strong enough to permit independent walk) | Yes / No | | | | | |
| 44 | Cognition - Qualitative evaluation | Normal / Abnormal | | | | | |
| 45 | Cognition – ECAS<br>observed value (normal range) | …………………/…………--………… | | | | | |
| 46 | Depression - Qualitative evaluation (Pessimism and loss of interest in activities once pleasurable or thoughts of suicide) | Normal / Abnormal | | | | | |
| 47 | Depression - ALS depression inventory<br>observed value /normal range | …………………/…………--………… | | | | | |
| | **5 – DISEASE SEVERITY AND PROGRESSION RATE** | | | | **Not feasible** | **Not available** | **Not relevant** |
| 48 | **Pattern of spreading**<br>Bulbar – 1<br>Right upper limb – 2<br>(2a-distal; 2b-proximal)<br>Left upper limb – 3<br>(3a-distal; 3b-proximal)<br>Right lower limb – 4<br>(4a-distal; 4b-proximal)<br>Left lower limb – 5<br>(5a-distal; 5b-proximal)<br>Respiratory symptoms – 6<br>Neck - 7<br>Thoracic /abdominal – 8<br>Dyscognition - 9<br><br>**Write progression** using → (e.g. 1→3a→6); if 2 regions are affected at the same time use **/** (e.g. 1→3a/6)<br>When not possible to define distal/prox just write the number (e.g. 3) | ……………………………………………………<br>……………………………………………………<br>……………………………………………………<br>……………………………………………………<br>…………………………………………………… | | | | | |
| 49 | **Timing of spreading from Region 1 to Region 2** (months) (Regions = Bulbar, Cervical, Lumbo-sacral, Thoracic, Dyscognition) | …………………………………………………… | | | | | |
| 50 | **Timing of spreading from Region 2 to Region 3** (months) (Regions = as above) | …………………………………………………… | | | | | |
| 51 | **ALSFRS-R** (current)<br>Bulbar subscore (nº 1+2+3) (B)<br>Upper limb subscore (nº 4+5+6) (UL)<br>Global subscore (nº 7) (G)<br>Lower limb subscore (nº 8+9) (LL)<br>Respiratory subscore (nº10+11+12) (R) | 1.<br>2.<br>3.<br>4. | 5.<br>6.<br>7.<br>8. | 9.<br>10.<br>11.<br>12. | | | |

| | | | | | |
|---|---|---|---|---|---|
| 52 | **ALSFRS-R rate of decay** = 48- current ALSFRSR /disease duration from first symptoms to current date (months) | | | | |
| | **6 – INVESTIGATIONS** (within 6 months before study entry) | | **Not feasible** | **Not available** | **Not relevant** |
| 53 | **CK** level /normal ranges (units) | ……………………/………………… (………………) | | | |
| 54 | **Albumin** level /normal ranges (units) | ……………………/………………… (………………) | | | |
| 55 | **Creatinine** level /normal ranges (units) | ……………………/………………… (………………) | | | |
| 56 | **Total Cholesterol** level /normal ranges (units) | ……………………/………………… (………………) | | | |
| 57 | **HDL Cholesterol** level /normal ranges (units) | ……………………/………………… (………………) | | | |
| 58 | **LDL Cholesterol** level /normal ranges (units) | ……………………/………………… (………………) | | | |
| 59 | **Triglycerides** level /normal ranges (units) | ……………………/………………… (………………) | | | |
| 60 | **Diagnostic EMG - bulbar region** | Normal / Abnormal (loss of motor units + fasciculation potentials and/or fibs-sw) | | | |
| 61 | **Diagnostic EMG - upper limbs** | Normal / Abnormal (loss of motor units + fasciculation potentials and/or fibs-sw) | | | |
| 62 | **Diagnostic EMG - lower limbs** | Normal / Abnormal (loss of motor units + fasciculation potentials and/or fibs-sw) | | | |
| 63 | **Diagnostic EMG - Nerve conduction studies - sensory neuropathy** | Yes / No | | | |
| 64 | **Brain MRI** | Normal (N) /Tumor (T) /Stroke (S) /Multiple sclerosis (MS) /Trauma (Tr) /Hypoxia (H) | | | |
| 65 | **Spinal cord MRI** Normal(N) /Tumor (T) / Stenosis(S) /Myelitis (My) /Trauma (Tr)/ Syringomyelia (S) / Other (O) | Cervical - …………………………………………<br>Thoracic - …………………………………………<br>Lumbo-sacral - ………………………………… | | | |
| 66 | **FVC** (% predicted values) | | | | |
| 67 | **VC** (% predicted values) | | | | |
| 68 | **Maximal Inspiratory Pressure** (% predicted values or value in cm $H_2O$) | | | | |
| 69 | **Maximal Expiratory Pressure** (% predicted values or value in cm $H_2O$) | | | | |
| 70 | **SNIP** – absolute value (cmH2O) | | | | |
| 71 | **Nocturnal oximetry** - mean value (%) | | | | |
| 72 | **Nocturnal oximetry < 90%** (% of the total time) | | | | |
| | **7 – CO-MORBIDITIES** (past and present) | | **Not feasible** | **Not available** | **Not relevant** |
| 73 | **Blood hypertension** | Yes / No | | | |
| 74 | **Diabetes – type I / II** | Yes (type 1 vs type 2) / No | | | |
| 75 | **Hypercholesterolemia** | Yes / No | | | |
| 76 | **Hypertriglyceridemia** | Yes / No | | | |
| 77 | **Hyperthyroidism** | Yes / No | | | |
| 78 | **Hypothyroidism** | Yes / No | | | |
| 79 | **Autoimmune rheumatologic disorder** | Yes / No | | | |
| 80 | **Autoimmune intestinal disorder** | Yes / No | | | |
| 81 | **Stroke** | Yes (ischemic vs hemorrhagic vs unknown) / No | | | |
| 82 | **Heart** – ischemia | Yes / No | | | |
| 83 | **Heart** – arrhytmia | Yes / No | | | |
| 84 | **Heart** – insufficiency | Yes / No | | | |

| | | | | | |
|---|---|---|---|---|---|
| 85 | **Cancer (primary)**<br>(multiple choice) | No / Yes:<br>Brain (B); Neck (N); Breast (Br); Lung (L); other Thoracic (T); Gastric (G); Colon (C); Rectum (Re); other Abdominal (A); Prostate (Pr); Uterus (U); other Pelvic (P); Spine (Sp); Bone cancer and sarcomas in upper limbs (UL); Bone cancer and sarcomas in lower limbs (LL); Skin (S); Leukemia (L); Lymphoma (Ly); other blood cancer (Bl). | | | |
| 85' | **Metastasis (above list )** | | | | |
| 86 | **Cancer treatment – Chemotherapy**<br>Specify the drug if possible | Yes / No<br>…………………………………………… | | | |
| 87 | **Cancer treatment – Radiotherapy**<br>**Head or neck** | Yes / No<br>Yes / No | | | |
| 88 | **Smoking**<br>Date of initiation (year)<br>Stopped<br>Date of interruption (year)<br>Mean number of packs/day | Yes / No<br>……………………………………………<br>Yes / No<br>……………………………………………<br>…………………………………………… | | | |
| | **8 – MEDICATION** (generic name) | | **Not feasible** | **Not available** | **Not relevant** |
| 89 | **Psychiatric** (> 6 months anytime)<br>Specify the drug if possible | Yes / No<br>…………………………………………… | | | |
| 90 | **Supplements** (> 6 months anytime)<br>Specify if possible | Yes / No<br>…………………………………………… | | | |
| 91 | **Riluzole**<br>Date of initiation<br>Side effects? | Yes / No<br>……………………………………………<br>…………………………………………… | | | |
| 92 | **Antiepileptc drug** (> 6 months anytime)<br>Specify if possible | Yes / No<br>…………………………………………… | | | |
| 93 | **Statins** (> 6 months anytime)<br>Specify if possible | Yes / No<br>…………………………………………… | | | |
| 94 | **NSAID** (> 6 months anytime)<br>Specify if possible | Yes / No<br>…………………………………………… | | | |
| 95 | **Steroids** (> 6 months anytime)<br>Specify if possible | Yes / No<br>…………………………………………… | | | |
| 96 | **Immunosupressors** (> 6 months anytime) Specify the drug if possible | Yes / No<br>…………………………………………… | | | |
| | **9 – GENETIC INFORMATION** | | **Not feasible** | **Not available** | **Not relevant** |
| 97 | **SOD1 mutation** (specify) | Yes……………………………………. / No | | | |
| 98 | **Abnormal C9orf72 repeat-expansion** | Yes / No | | | |
| 99 | **Other genes**<br>**Was the patient included in genomic studies** (specify) | Yes / No<br>Yes……………………………………. / No | | | |
| 100 | **Familial history of ALS** (Multiple choice) | No<br>mother<br>father<br>heterozygous twin brother<br>homozygous twin brother<br>heterozygous twin sister<br>homozygous twin sister<br>Other family members:<br>……………………………………………………………<br>……………………………………………………………<br>……………………………………………………………<br>…………………………………………………………… | | | |

| | | |
|---|---|---|
| 101 | **Familial history of FTD** | No<br>mother<br>father<br>heterozygous twin brother<br>homozygous twin brother<br>heterozygous twin sister<br>homozygous twin sister<br>Other family members:<br>……………………………………………………………………<br>……………………………………………………………………<br>……………………………………………………………………<br>……………………………………………………………………<br>……………………………………………………………………<br>…………………………………………………………………… |
| 102 | **Familial history of Alzheimer disease** | No<br>mother<br>father<br>heterozygous twin brother<br>homozygous twin brother<br>heterozygous twin sister<br>homozygous twin sister<br>Other family members:<br>……………………………………………………………………<br>……………………………………………………………………<br>……………………………………………………………………<br>……………………………………………………………………<br>……………………………………………………………………<br>…………………………………………………………………… |
| 103 | **Familial history of Parkinson disease** (which family member) | No<br>mother<br>father<br>heterozygous twin brother<br>homozygous twin brother<br>heterozygous twin sister<br>homozygous twin sister<br>Other family members:<br>……………………………………………………………………<br>……………………………………………………………………<br>……………………………………………………………………<br>……………………………………………………………………<br>……………………………………………………………………<br>…………………………………………………………………… |
| 104 | **Familial history of multiple sclerosis** (which family member) | No<br>mother<br>father<br>heterozygous twin brother<br>homozygous twin brother<br>heterozygous twin sister<br>homozygous twin sister<br>Other family members:<br>……………………………………………………………………<br>……………………………………………………………………<br>……………………………………………………………………<br>……………………………………………………………………<br>……………………………………………………………………<br>…………………………………………………………………… |
| 105 | **Other severe diseases in the family** | 1. Mother……………………………………………………<br>……………………………………………………………………<br>……………………………………………………………………<br>……………………………………………………………………<br>2. Father……………………………………………………<br>…………………………………………………………………… |

| | | | | | | |
|---|---|---|---|---|---|---|
| | | ………………………………………………………………………<br>3. ……………………………………………………………………<br>4. ……………………………………………………………………<br>5. ……………………………………………………………………<br>6. ……………………………………………………………………<br>7. ……………………………………………………………………<br>8. ……………………………………………………………………<br>9. ……………………………………………………………………<br>10.…………………………………………………………………… | | | | |
| 106 | **Parents' and siblings´ causes of death** | 1. Mother: Alive Yes / No<br>……………………………………………………<br>2. Father: Alive Yes / No<br>……………………………………………………<br>3. ……………………………………………<br>4. ……………………………………………<br>5. ……………………………………………<br>6. ……………………………………………<br>7. ……………………………………………<br>8. ……………………………………………<br>9. ……………………………………………<br>10. ………………………………………… | Age<br>………..<br><br>………..<br>………..<br>………..<br>………..<br>………..<br>………..<br>………..<br>………..<br>……….. | | | |
| 107 | **Mother´s number of siblings and birth order** (e.g. 2nd child in 7, should be 2/7) | | | | | |
| 108 | **Father´s number of siblings and birth order** (e.g. 2nd child in 7, should be 2/7) | | | | | |
| 109 | **Consanguinity of parents** | Yes / No | | | | |
| 110 | **Number of Siblings and birth order** (e.g. 2nd child in 7, should be 2/7)<br>With monozygous twin<br>With a dizygous twin | ……………………………………………….<br>Yes / No<br>Yes / No | | | | |
| | **10 – HABITS / TRAUMA AND SURGERY** | | | **Not feasible** | **Not available** | **Not relevant** |
| 111 | **Regular physical exercise** | Yes / No | | | | |
| 112 | **Intense physical exercise** > 1 year<br><br>1: 150 min/week of moderate aerobic activity (such as fast walking, swimming) | **Which sport:**<br>Basketball (BA); Boxing(BO); Climbing (CL); Cycling (CY); Dancing (D); Diving (DV); Fast walking (FW); Fencing (FE); Fitness (F); Football (FB); Golf (G); Handball (HB); Hockey (HO); Rowing (RO); Rugby (RU); Running (R); Skiing (SK); Swimming (S); Tennis (T); Volleyball (V); Weightlifting(WL)<br>Others: | **How long (yrs):**<br>e.g.: DV: 10, SK 25 (diving 10 yrs, skiing 25 yrs) | | | |
| | 2: 75 min/week of vigorous aerobic activity (such as running or aerobic dancing) | **Which sport:**<br>Basketball (BA); Boxing(BO); Climbing (CL); Cycling (CY); Dancing (D); Diving (DV); Fast walking (FW); Fencing (FE); Fitness (F); Football (FB); Golf (G); Handball (HB); Hockey (HO); Rowing (RO); Rugby (RU); Running (R); Skiing (SK); Swimming (S); Tennis (T); Volleyball (V); Weightlifting(WL)<br>Others: | **How long (yrs):** | | | |

| | | | |
|---|---|---|---|
| 113 | **Mild physical exercise** > 1 year<br>(< intensity above) | **Which sport:**<br>Basketball (BA); Boxing(BO); Climbing (CL); Cycling (CY); Dancing (D); Diving (DV); Fast walking (FW); Fencing (FE); Fitness (F); Football (FB); Golf (G); Handball (HB); Hockey (HO); Rowing (RO); Rugby (RU); Running (R); Skiing (SK); Swimming (S); Tennis (T); Volleyball (V); Weightlifting(WL)<br>Others: | **How long (yrs):** |
| 114 | **Regular specific diet** > 1 year (specify) | No<br>Mediterranean (M); Vegetarian (VT); Vegan (VG); Gluten-free (GF); High protein (HP); Low carbohydrate (LC); Paleo (High- protein, high-fat low-carbohydrate) (PA) | |
| 115 | **Head/Neck trauma** in the last 5 years before disease onset (only if hospitalized) | Yes (head vs neck) / No | |
| 116 | **Head/Neck trauma** > 5 years before disease onset (only if patient was hospitalized) | Yes (head vs neck) / No | |
| 117 | **Other trauma** in the last 5 years before disease onset (only if hospitalized)<br>Cervical (C)<br>Thoracic (T)<br>Lumbo-sacral (LS) | Yes (C, T, L-S) / No | |
| 118 | **Other trauma** > 5 years before disease onset (only if patient was hospitalized)<br>Cervical (C)<br>Thoracic (T)<br>Lumbo-sacral (LS) | Yes (C, T, L-S) / No | |
| 119 | **Spine surgery** in the last 5 years before disease onset<br>Cervical (C)<br>Thoracic (T)<br>Lumbo-sacral (LS) | Yes (C, T, L-S) / No | |
| 120 | **Spine surgery** > 5 years before disease onset<br>Cervical (C)<br>Thoracic (T)<br>Lumbo-sacral (LS) | Yes (C, T, L-S) / No | |
| 121 | **Limb surgery**<br>in the last 5 years before disease onset<br>Upper limb (UL)<br>Lower lib (LL) | Yes (UL, LL) / No | |
| 122 | **Limb surgery** > 5 years before disease onset<br>Upper limb (UL)<br>Lower limb (LL) | Yes (UL, LL) / No | |
| 123 | **Abdominal surgery** in the last 5 years before disease onset | Yes / No | |
| 124 | **Abdominal surgery**<br>> 5 years before disease onset | Yes / No | |
| 125 | **Thoracic surgery**<br>in the last 5 years before disease onset | Yes / No | |
| 126 | **Thoracic surgery**<br>> 5 years before disease onset | Yes / No | |

| | | | | | |
|---|---|---|---|---|---|
| 127 | **Pelvic Surgery**<br>in the last 5 years before disease onset | Yes / No | | | |
| 128 | **Pelvic Surgery**<br>> 5 years before disease onset | Yes / No | | | |
| 129 | **Head/neck Surgery**<br>in the last 5 years before disease onset | Yes / No | | | |
| 130 | **Head/neck Surgery**<br>> 5 years before disease onset | Yes / No | | | |
| 131 | **Blood transfusion**<br>in the last 5 years before disease onset | Yes / No | | | |
| 132 | **Blood transfusion**<br>> 5 years before disease onset | Yes / No | | | |
| 133 | **Blood donor**<br>in the last 5 years before disease onset | Yes / No | | | |
| 134 | **Blood donor**<br>> 5 years before disease onset | Yes / No | | | |
| 135 | **Electric shock** in the last 5 years before disease onset (if patient was hospitalized) | Yes / No | | | |
| 136 | **Electric shock** > 5 years before disease onset (leading to hospitalization of patient) | Yes / No | | | |
| 137 | **Emotional stress** in the last 5 years before disease onset (loss or disease of close family member, loss of job ) | Yes / No | | | |
| 138 | **Emotional stress** >5 years before disease onset (loss or disease of close family member, loss of job ) | Yes / No | | | |
| | **11 – OCCUPATIONS** | | **Not feasible** | **Not available** | **Not relevant** |
| 139 | **Main Occupation**<br>in the last 5 years before disease onset | | | | |
| 140 | **Main Occupation**<br>>5 years before disease onset | | | | |
| 141 | **Second Occupation**<br>in the last 5 years before disease onset | | | | |
| 142 | **Second Occupation**<br>> 5 years before disease onset | | | | |
| 143 | **Place of living**<br>in the last 5 years before disease onset | …………………………/……………………….<br>Village - < 1000 inhabitants<br>Country towns– 1000-5 000<br>Small town– 5 000-20 000<br>Middle town– 20 000 – 100 000<br>Large town– > 100 000 | | | |
| 143’ | **Place of living characteristics**<br>in the last 5 years before disease onset<br>**(multiple choice)** | Rural area (RA); Urban area (UA);<br>Near sewage plant (NSP);<br>Near landfills (NL);<br>Near waste incinerators (MWI);<br>Near sources of EM fields (high-voltage transmission lines, radar etc.) (EMF) | | | |
| 144 | **Place of living**<br>> 5 years before disease onset | …………………………/……………………….<br>Village - < 1000 inhabitants<br>Country towns– 1000-5 000<br>Small town– 5 000-20 000<br>Middle town– 20 000 – 100 000<br>Large town– > 100 000 | | | |
| 144’ | **Place of living characteristics**<br>> 5 years before disease onset<br>**(multiple choice)** | Rural area (RA); Urban area (UA);<br>Near sewage plant (NSP);<br>Near landfills (NL);<br>Near waste incinerators (MWI);<br>Near sources of EM fields (high-voltage transmission lines, radar etc.) (EMF) | | | |

| | | |
|---|---|---|
| 145 | **ALS cases in the neighborhood** | Yes / No |
| 146 | **ALS disease in co-workers** | Yes / No |
| 147 | **ALS disease in friends** | Yes / No |
| 148 | **First specialist who observed the patient** | |
| 149 | **Investigation requested by the first specialist:**<br>Brain CT scan<br>Cervical spinal cord CT scan<br>Lumbo-sacral spinal cord CT scan<br>Brain MRI<br>Cervical spinal cord MRI<br>Thoracic spinal cord MRI<br>Lumbo-sacral spinal cord MRI<br>EMG | <br><br>Yes / No<br>Yes / No<br>Yes / No<br>Yes / No<br>Yes / No<br>Yes / No<br>Yes / No<br>Yes / No |
| 150 | **Second specialist who observed the patient** | |
| 151 | **Time gap between first-second specialist** (months) | |
| 152 | **Investigation requested by the second specialist:**<br>Brain CT scan<br>Cervical spinal cord CT scan<br>Lumbo-sacral spinal cord CT scan<br>Brain MRI<br>Cervical spinal cord MRI<br>Thoracic spinal cord MRI<br>Lumbo-sacral spinal cord MRI<br>EMG | <br><br>Yes / No<br>Yes / No<br>Yes / No<br>Yes / No<br>Yes / No<br>Yes / No<br>Yes / No<br>Yes / No |
| 153 | **Specialist who made the diagnosis** | |
| 154 | **Order of specialists that observed the patient**<br>First (specialty) /made the diagnosis?<br>Second (specialty) /made the diagnosis?<br>Third (specialty) /made the diagnosis?<br>Fourth (specialty) /made the diagnosis?<br>Fifth (specialty) /made the diagnosis?<br>Other (specialty)/made the diagnosis? | <br><br>………………………………………… Yes / No<br>………………………………………… Yes / No<br>………………………………………… Yes / No<br>………………………………………… Yes / No<br>………………………………………… Yes / No<br>………………………………………… Yes / No |
| 155 | **Time gap between first medical observation and the specialist who made the diagnosis** (months) | |
| 156 | **Investigation requested by the specialist who made the diagnosis:**<br>Brain CT scan<br>Cervical spinal cord CT scan<br>Lumbo-sacral spinal cord CT scan<br>Brain MRI<br>Cervical spinal cord MRI<br>Thoracic spinal cord MRI<br>Lumbo-sacral spinal cord MRI<br>EMG | <br><br>Yes / No<br>Yes / No<br>Yes / No<br>Yes / No<br>Yes / No<br>Yes / No<br>Yes / No<br>Yes / No |
